# Applying the Analytical Hierarchy Process to global health decision making: Active pharmaceutical ingredient – technology (API-tech) mapping case studies, lessons learned, and opportunities

**DOI:** 10.1101/2025.08.27.25334532

**Authors:** Caroline R Soyars-Tetarbe, Hal Forman, Melissa J Leavitt, Melynda Watkins, Paul Domanico, Ernest Forman

**Author notes:** Corresponding author: (CST). These authors contributed equally to this work (Caroline R Soyars-Tetarbe, Hal Forman, Melissa J Leavitt). These authors also contributed equally to this work (Melynda Watkins, Paul Domanico).

## Abstract

The Analytical Hierarchy Process (AHP) is a well-established decision science tool that has been widely applied across various sectors. Despite the pervasiveness of AHP across many industries, there is limited documented evidence of this methodology being applied to global health contexts. The purpose of this study was to demonstrate how AHP can be applied to global health challenges, specifically to prioritize investments in pediatric antibiotics and antiretrovirals. The Expert Choice Comparion^TM^ software was used to build AHP surveys to achieve the goal of prioritizing age-appropriate formulations according to how effectively they meet the desired product attributes. The AHP surveys were disseminated to community members, clinicians, and programmatic experts to provide input on the relative importance of the product attributes and products by responding to pairwise and Likert-based questions. Results were subsequently analyzed and interpreted using Expert Choice. Results from the pediatric antibiotics AHP survey revealed that standard oral dosage forms, microarray patches, and oral films were similarly high priority across pediatric sub-populations. For the pediatric HIV AHP survey, oral films, injectables, and microarray patches were top scoring platforms across pediatric sub-populations, with implant-depot formulations also scoring highly for older children. The AHP findings were used to develop product-specific roadmaps and calls to action for how donors, developers, and other key stakeholders should prioritize investments to accelerate development of the most promising technologies. The successful utilization of AHP in this study provides an evidence base on applying this methodology to define investment priorities in global health settings. These findings suggest that there may be an opportunity for broader use of AHP in global health to support resource allocation, investment prioritization, and program implementation amidst an uncertain and constrained global health funding landscape.

## Introduction

Multi criteria decision making methods have been adopted by decision makers to support initiatives focused on achieving targets outlined in the UN Sustainable Development Goals (SDGs) [1]. However, only 17% of SDG targets are on track to be reached by the 2030 deadline including slowed, stalled, or reversed progress towards targets within SDG 3 which pertains to health and wellbeing [2]. There may be an opportunity to further improve, optimize, and implement decision science tools to help navigate pain points that are impeding progress of the SDGs.

The depletion of global health funding in recent years heightens the need for robust, science-based methods for prioritizing investments. Some forecasts estimate that global health assistance could be reduced by approximately 50% by 2030 in favor of being reallocated to competing funding priorities such as addressing impacts of climate change [3]. The dismantling of the United States Agency for International Development (USAID) has introduced additional uncertainties, halting US contributions to The Global Fund and the President’s Emergency Plan for AIDS Relief (PEPFAR) which could lead to more than 14M additional deaths within the same time frame [4]. These significant shifts exacerbate the opportunity to explore frameworks that could assist with prioritizing investments in extremely constrained funding environments, and ultimately support efforts to ensure that the significant gains in reducing mortality in low- and middle-income countries (LMICs) over the last few decades are not reversed.

The Analytical Hierarchy Process (AHP) is a multicriteria decision science tool used by individuals, groups, and organizations to support complex decision making [5]. Through AHP, these groups can assess how well a list of alternative solutions achieve a goal by how well they address contributing factors for the goal. AHP uses pairwise comparisons to break down larger, complex decisions into multiple, pair-wise decisions, thereby enabling both qualitative and quantitative factors to be systematically considered. The AHP framework has been used widely across multiple industries including business, healthcare, engineering, government policy, and project portfolio management [6–9].

Global health decision-making at all levels must consider numerous complex factors such as sociocultural, financial, technical, clinical, environmental, legal, and political topics. Despite the inherent complexity of global health decisions and the use of these tools in high-income settings, there is limited published evidence on applying AHP to programs focused on these challenges in LMICs [1, 10, 11]. The lack of documented examples of AHP use in LMIC healthcare contexts presents an opportunity to explore how AHP can be applied to tackle the highest priority health needs faced by underserved communities around the globe.

The authors have now applied AHP to multiple high priority global health challenges, such as age-appropriate medications for pediatric populations. The purpose of this paper is to present findings from case studies that focus on the implementation of the AHP framework to prioritize the development of pediatric antibiotic and human immunodeficiency virus (HIV) formulations for LMICs using the Expert Choice Comparion^TM^ software. The results from these case studies demonstrate the power of AHP to define investment priorities and generate actionable recommendations when applied to a global health setting. These findings uncover the opportunity for broader uptake of the AHP framework within the global health community to ensure health gains over the last few decades are not lost as funding for global health shrinks.

## Materials and methods

### Ethics statement

The study did not involve human research participants or human research participants’ data and therefore is exempt from requiring ethics approval.

### AHP

AHP is a widely used structured decision-making framework, particularly prominent in scientific research across the globe [6, 7, 12]. AHP is applied across various fields to support complex decision-making. In business and management, it is used for tasks such as supplier selection, project prioritization, and developing marketing strategies [13]. In engineering, AHP assists in decisions like material selection and site selection [14]. The healthcare sector utilizes AHP for treatment planning and determining optimal hospital locations [15]. In environmental science, it supports decisions related to renewable energy project selection and waste management strategies [16]. In the field of education, AHP is applied to curriculum design, university rankings, and guiding scientific research [17]. Lastly, in information technology, it helps with software selection and the development of cybersecurity strategies [18].

One of the core mathematical foundations of AHP involves the application of eigenvectors and eigenvalues, which are used to derive precise ratio-scale priorities from pairwise comparison matrices [19–21]. This mathematical approach ensures that the priorities generated are both consistent and reflective of the decision maker’s judgments. AHP facilitates the decomposition of multifaceted decision problems into a hierarchical structure, enabling systematic analysis while integrating inputs from multiple stakeholders to capture diverse evaluative criteria [6, 8, 12, 22].

AHP integrates both qualitative and quantitative criteria by utilizing ratio-scale numbers, which allows for precise and consistent comparisons [13, 23]. One of its key strengths is its ability to facilitate rational judgments, thereby reducing bias and ensuring logical consistency in decision-making. AHP also enables sensitivity analysis, allowing users to test how changes in the weights of criteria can influence the final prioritization of alternatives [24, 25]. Furthermore, AHP is well suited for group decision-making, as it aggregates the judgments of multiple stakeholders and subject-matter experts. This collaborative approach not only enhances the quality of the decision but also fosters greater buy-in and satisfaction among participants [26].

Decision science methodologies, including AHP, are valuable for overcoming a range of cognitive biases that often hinder effective decision-making, including:

- Limited memory capacity [27]
- Framing bias [28, 29]
- Overconfidence [30]
- Politics within organizations [31]
- Groupthink [32, 33]
- Loss aversion [34]
- Sunk cost fallacy [35, 36]
- Endowment effect [37]
- Availability bias and anchoring [38–40]
- Representativeness bias [41]

AHP also helps avoid several common and often counterproductive decision-making strategies, including:

- Reliance on intuition [42–44]
- Conference room paralysis
- Rules of thumb
- Heuristics
- Nutshell briefings
- Satisficing
- Misuse of numbers

The original AHP theory had certain limitations, one of the most notable being the rank reversal problem [45]. This issue arises when the addition or removal of an alternative unexpectedly alters the ranking of the remaining options. To overcome this, the Ideal Synthesis method was introduced in 1993 [22]. This approach addresses rank reversal by ensuring that the relative rankings of existing alternatives remain stable, regardless of changes to the set of options.

Another criticism of the Analytic Hierarchy Process (AHP) is the time required to perform the pairwise comparisons. These comparisons are processed using a powerful mathematical technique known as the principal right-hand eigenvector, which effectively reduces errors when translating human judgment into ratio-level measures of the decision makers’ subjective prioritization of objectives. The hierarchical structuring inherent in AHP helps minimize the number of necessary comparisons, making it suitable for decisions involving a small number of alternatives. For decisions with many alternatives, a “ratings” technique—introduced to AHP years ago—further reduces the number of required comparisons.

Although the comparison process can be time-consuming, tasks are typically distributed among participants based on their expertise and roles. Moreover, the logical structure of AHP often accelerates decision-making by reducing repetitive debate. The time devoted to AHP might also be recovered by efficiencies elsewhere.

### Expert Choice software

The Expert Choice software (https://www.expertchoice.com/comparion), is a structured decision-making method that uses AHP to evaluate and prioritize options based on multiple criteria. It involves breaking down a complex decision into a hierarchy, starting with the overall goal at the top, followed by the criteria and sub-criteria that influence the decision, and finally the available alternatives. Within this framework, decision makers perform pairwise comparisons between elements at each level of the hierarchy—such as comparing how much more important one criterion is over another. These comparisons are typically made using the fundamental verbal AHP ordinal scale ranging from 1 to 9.

While these values represent ordinal judgments rather than true ratio scale measurements, applying an eigenvector computation helps convert them into ratio scale values. This process minimizes the errors typically associated with ordinal data and enhances the accuracy of the resulting priorities. The software then calculates numerical weights that reflect the relative importance of each element, helping users rank the alternatives in a rational and transparent way. Expert Choice is especially useful in group settings, as it facilitates consensus and ensures that all perspectives are considered systematically.

Expert Choice offers additional types of platforms to support different types of decisions. One of the standout features across Expert Choice platforms is an emphasis on team collaboration and transparency. The platform includes built-in tools for communication, group assessments, and decision documentation, which enable enhanced understanding, faster processes, and improved alignment across stakeholders.

### API-tech mapping

The case studies discussed in this paper, hereby referred to as API-tech mapping, was developed by CHAI in collaboration with the World Health Organization (WHO) and its Global Accelerator for Paediatric Formulations Network (GAP-f) partners to prioritize the most impactful age-appropriate medications for pediatric populations. The purpose of API-tech mapping exercises is to systematically identify and prioritize the pairing of active pharmaceutical ingredients (API) to drug delivery mechanisms (‘tech’) that hold the greatest potential for investment. This prioritization process serves multiple functions, including helping to build consensus among stakeholders, setting a clear research and development agenda, and ensuring that resources are directed towards the most impactful solutions.

API-tech mapping exercises are guided by decision science methodologies, specifically AHP, and involve a multi-faceted, stakeholder-driven approach. The process consists of seven main steps, which are summarized in Fig 1. Step 5 (clinical assessment) is the focus of the case studies in this paper since this is the step that utilizes the AHP framework.

**Fig 1.**
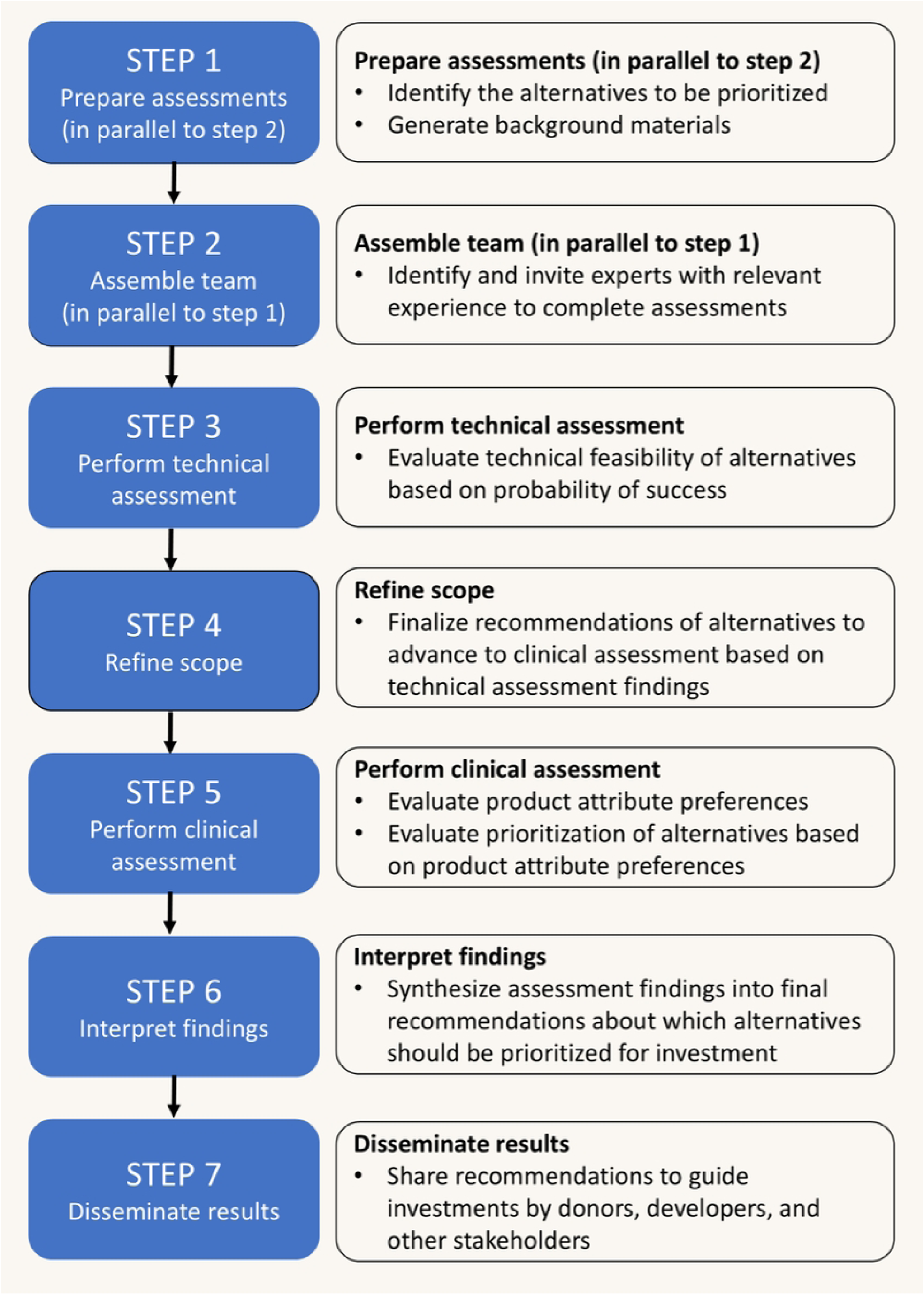
Overview of API-tech mapping process.

### Case studies: Leveraging AHP to prioritize pediatric drug development

Improving the health and well-being of newborns, children, and adolescents around the world has been a long-standing priority of WHO, GAP-f [46], and other critical global health actors. WHO leads multiple ongoing initiatives such as the WHO Model List of Essential Medicines for Children [47] and Pediatric Drug Optimization (PADO) process [48] to improve access to safe, effective, and age-appropriate medicines for children globally.

Although increased access to pediatric medicines has been identified as a critical global health priority, the private sector has been slow to respond to this gap. Children are a diverse and dynamic population, and as a result, drug developers have historically struggled with developing formulations that meet pediatric needs whilst achieving the desired therapeutic effect. For example, it is widely understood that poor-tasting pediatric oral medications are a barrier to compliance [49–51]. In addition to technical and user-centric challenges, pediatric populations represent a small segment of the pharmaceutical market. As a result, developers are oftentimes disincentivized from making significant investments into the research and development of alternative drug delivery platforms tailored to the unique needs of pediatric patients.

In response to this identified need and in alignment with PADO priorities, WHO partnered with CHAI to conduct API-tech mapping exercises to prioritize pediatric antibiotics and HIV formulations for LMICs. The AHP exercises conducted within the broader API-tech mapping process are described as two independent case studies below.

## Case study 1: Prioritization of pediatric antibiotic formulations for LMICs

### Objectives and scope

An AHP exercise was launched with the goal of prioritizing the compatibility and likelihood of product development success for the most promising innovative technologies when compared with traditional standard solid oral dosage forms in the context of pediatric antibiotics for shorter term, systemic delivery. The APIs, products, and product attributes that were used as the AHP framework inputs are in **Table 1. Summary of APIs (top left), products (top right), and product attributes (bottom) in scope for the AHP pediatric antibiotics exercise.**Table 1. This scope was defined based on the PADO antibiotics report outputs [52], technical assessments, and core team engagements prior to initiating the AHP assessment.

**Table 1.**
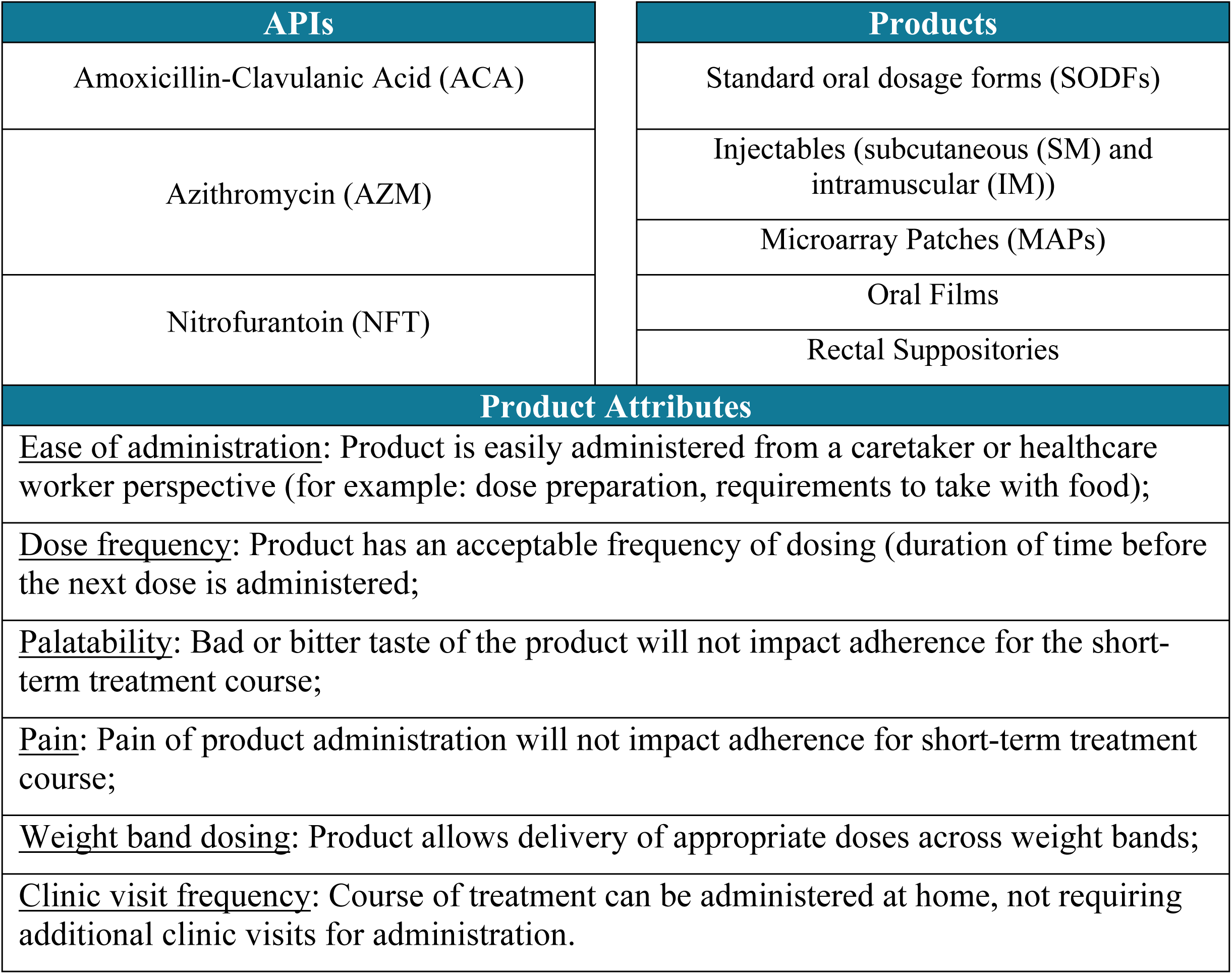
Summary of APIs (top left), products (top right), and product attributes (bottom) in scope for the AHP pediatric antibiotics exercise.

| APIs | Products |
| --- | --- |
| Amoxicillin-Clavulanic Acid (ACA) | Standard oral dosage forms (SODFs) |
| Azithromycin (AZM) | Injectables (subcutaneous (SM) and intramuscular (IM)) |
|  | Microarray Patches (MAPs) |
| Nitrofurantoin (NFT) | Oral Films |
|  | Rectal Suppositories |
| Product Attributes |  |
| <u>Ease of administration</u>: Product is easily administered from a caretaker or healthcare worker perspective (for example: dose preparation, requirements to take with food); |  |
| <u>Dose frequency</u>: Product has an acceptable frequency of dosing (duration of time before the next dose is administered); |  |
| <u>Palatability</u>: Bad or bitter taste of the product will not impact adherence for the short-term treatment course; |  |
| <u>Pain</u>: Pain of product administration will not impact adherence for short-term treatment course; |  |
| <u>Weight band dosing</u>: Product allows delivery of appropriate doses across weight bands; |  |
| <u>Clinic visit frequency</u>: Course of treatment can be administered at home, not requiring additional clinic visits for administration. |  |

### AHP model

Once the inputs were established, they were entered into the Expert Choice software to automatically generate a survey based on the AHP framework. The Expert Choice survey was divided into two parts: 1) Derive the relative importance for each desired product attribute in achieving the overall goal; and 2) Determine the priority of products by their ability to achieve the desired product attributes.

Each part was repeated for each of the three pediatric sub-populations that were defined for this specific exercise – neonates and infants (0 up to 2 years), younger children (2 years up to 5 years), older children (5 years up to 12 years). Adolescents over 12 years old were excluded because the assumption was that these children would take adult dosing and therefore are not critical for understanding administration challenges and limitations. The exclusion of this age group is also aligned with the scope of the WHO Essential Medicines list for children.

The first part of the Expert Choice survey to derive the relative importance for each desired product attribute was achieved by a series of pairwise questions to compare the relative importance of the six desired product attributes. The objectives hierarchy feature of the Expert Choice software was used to structure the decision model such that data could be collected and segmented by pediatric sub-populations which were hypothesized to have different preferences.

Fig 2 shows an example of a pairwise comparison of the desired product attributes for ease of administration and pain specifically for the sub-population of neonates and infants (0 up to 2 years). Using the scale provided, respondents select the bar that represents how they compare the importance of the two product attributes listed. The same pairwise format was used to compare the other desired product attributes, so that each of the six desired product attributes are compared against every other attribute.

**Fig 2.**
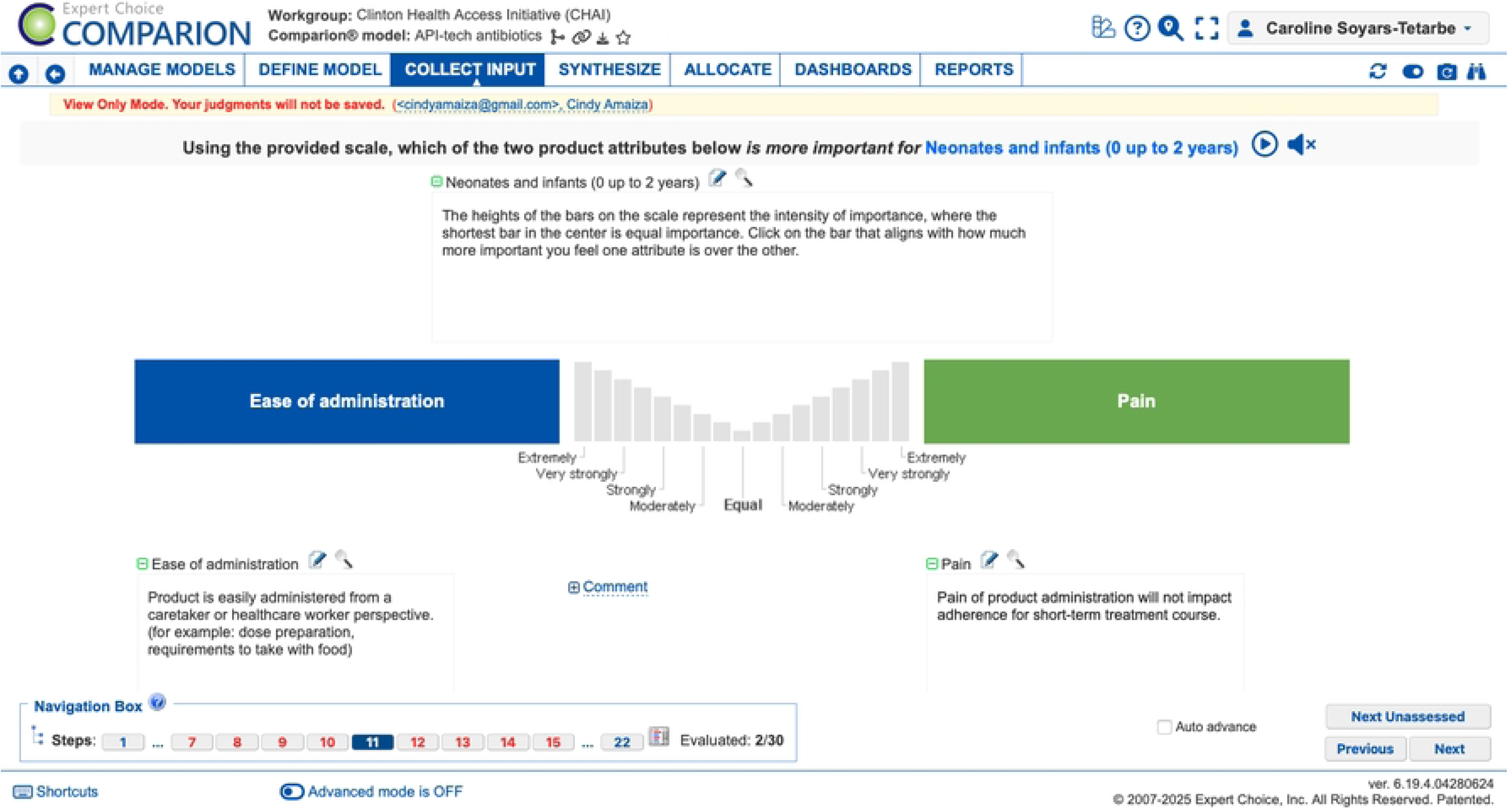
Example of a pairwise comparison question from the Expert Choice survey to derive the relative importance for each desired product attribute for each pediatric sub population.

The second part of the survey to prioritize the alternative products was achieved by a series of questions asking the respondent to agree or disagree on a 5 point Likert scale [53] on whether each product achieved the desired product attribute. Fig 3 shows an example of a Likert scale question for rating how well each product achieves the desired product attribute of the product being easy to administer, specifically for the sub-population of neonates and infants (0 up to 2 years).

**Fig 3.**
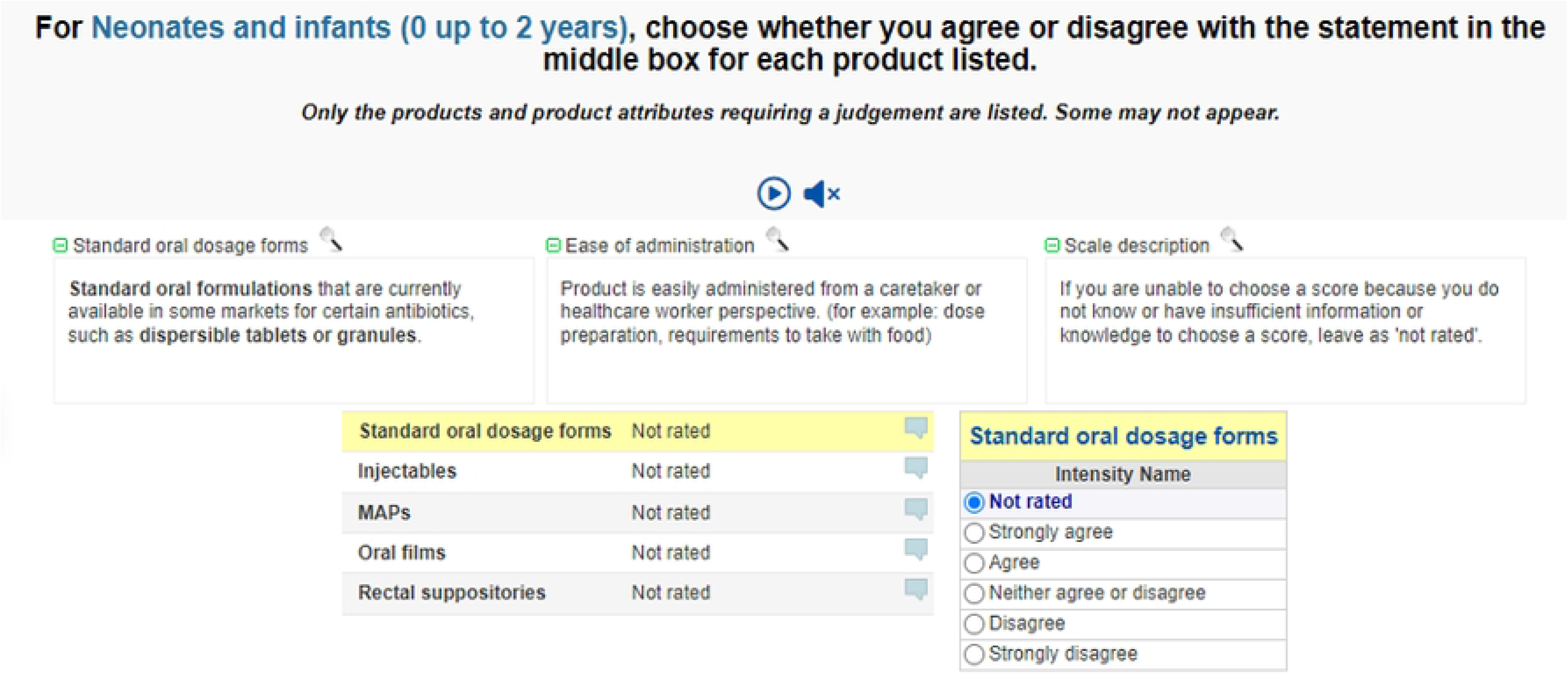
Example of a question from the Expert Choice survey to prioritize how well each product achieves the desired product attribute.

Using the Likert scale, respondents select how much they agree or disagree with the desired product attribute listed in the upper middle section. This same format was used to rate each product against each desired product attribute within the three pediatric sub-populations.

The Contributions function (Fig 4) within the Expert Choice software was used to reduce the number of judgements completed by the respondent. Broadly speaking, this function allows the user to specify which objectives (in this case, the product attributes) are applicable to each of the alternatives (in this case, the products). For example, palatability is not applicable for injectables, MAPs, or rectal suppositories since those are not oral formulations. This feature allowed the team to only select criterion that were applicable to each product, thus reducing the length of the survey.

**Fig 4.**
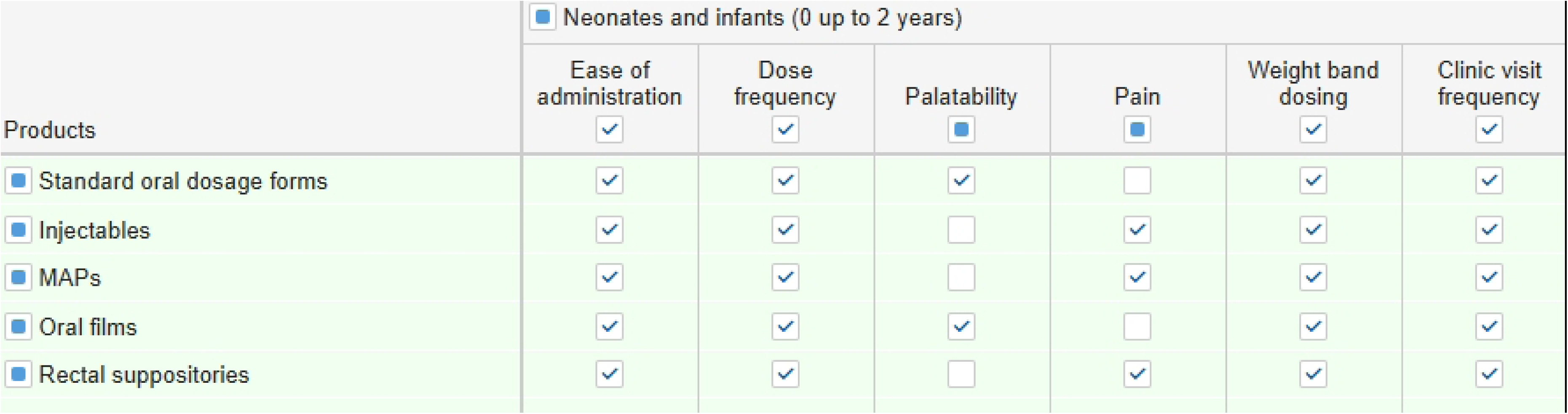
Contributions function in the Expert Choice software to select combinations of products and product attributes to include in the survey.

### AHP analysis and results

The tool allowed the team to survey a wide array of global stakeholders by providing survey responses via a unique link. Of the 81 global stakeholders invited to participate in the pediatric antibiotics survey, 28 people responded across clinical, regulatory/normative, and community member stakeholder groups. Depending on the number of responses, the Expert Choice software has the capability to segment the data based on pre-determined demographics such as location or stakeholder group. For this case study, the data was pooled across stakeholder groups for analyses due to a small sample size despite multiple attempts to get additional and diverse responses.

The Expert Choice software algorithm and data visualization features were utilized to analyze and interpret results. For each pediatric sub-population, the Expert Choice software aggregated the data, performed calculations, and created visualizations of the relative importance for the desired product attributes and the prioritization of products overall and by desired product attribute. Fig 5 and Fig **6**.

**Fig 5.**
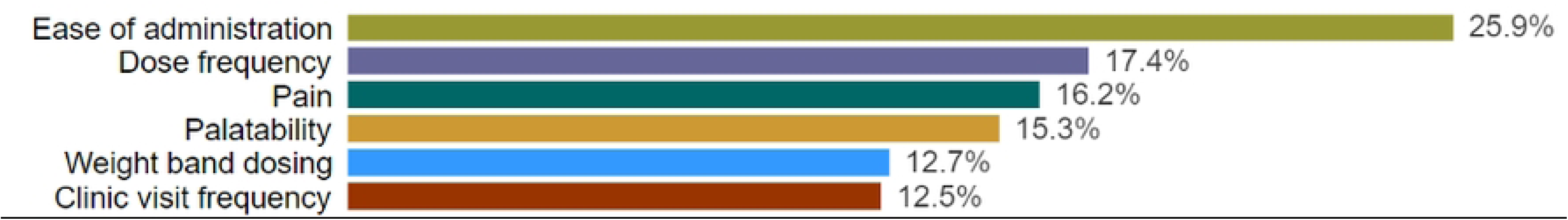
Pooled results of the order of importance for each desired product attribute within the neonates and infants age group to illustrate data analyses and visualization for product attributes. Each bar indicates a desired product attribute, and the priority is listed to the right of the bar.

**Fig 6.**
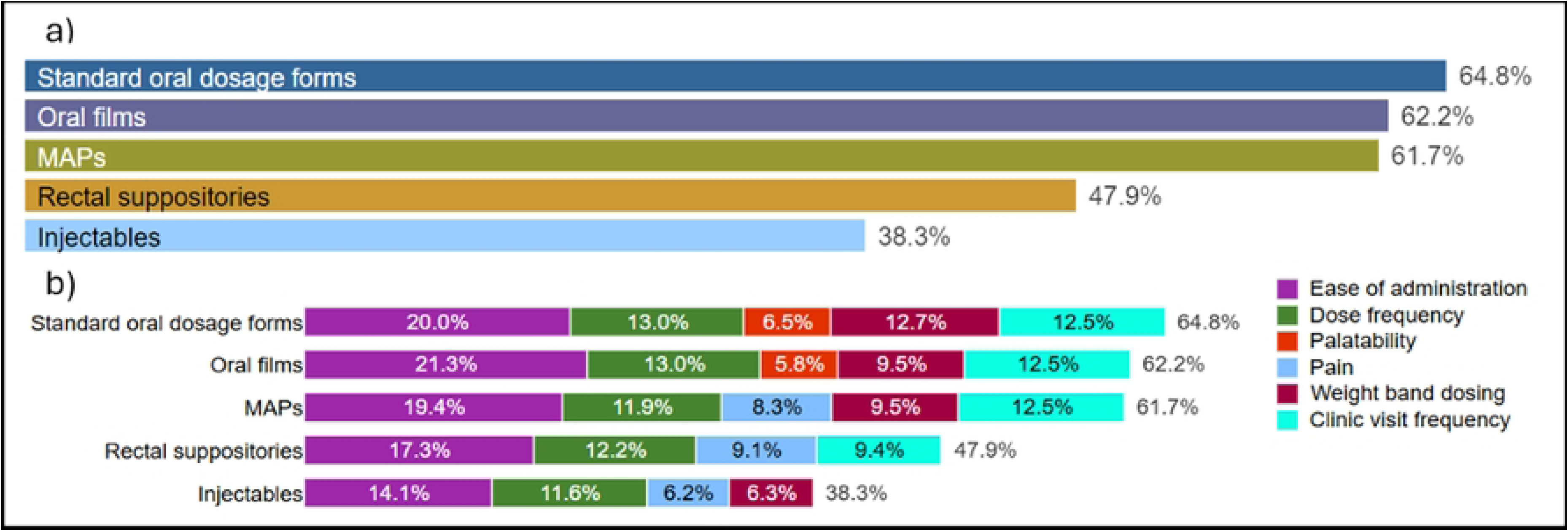
Pooled, unnormalized results of product prioritization for neonates and infants age group to illustrate data analysis and visualization for product prioritization. Figure 6a and 6b are without and with the product attribute breakdown for each product, respectively. Note that pain is not applicable to SODFs and oral films, and palatability is not applicable to MAPs.

**Pooled, unnormalized results of product prioritization for neonates and infants age group to illustrate data analysis and visualization for product prioritization.** Figureprovides an illustrative example of the data visualizations that were generated by the Expert Choice software for each pediatric sub-population and used to interpret results. Fig 5 results are presented as normalized values, where the summation of the six product attribute scores is 100%. Figs Fig 6. Pooled, **unnormalized results of product prioritization for neonates and infants age group to illustrate data analysis and visualization for product prioritization.** Figureresults are presented as unnormalized values, which represent the total score for the product out of a possible 100%, where 100% would be scoring the highest possible score across all desired product attributes. Fig 6b illustrates how a user can look at the overall scores or break down the total score by each criterion in case there are differences that warrant further interrogation. For example, the data presented in Figs 5 and 6 leads to the conclusion that ease of administration is the single most important product attribute for neonatal and infant populations and the high priority products for this age group are SODFs, oral films, and MAPs.

As part of the Expert Choice software, users can utilize the Dynamic Analysis feature to perform sensitivity analyses. The Dynamic Analysis feature allows the user to drag the objective priorities back and forth in the left column and see how the alternatives (in this case, the products) are impacted, ultimately visualizing how sensitive the objectives and alternatives are to each other. Fig 7 shows an example of how this sensitivity analysis was performed for this case study. The grey vertical marker indicates the original value for each bar, and by sliding the bar of an attribute, you can see the impact on both the weightings of the other attributes, which increase depending on their weighting, and the impact on the product prioritization. For this case study, the conclusions and product priorities did not change significantly. In some cases, there were some noticeable shifts to prioritization, however, the top prioritized products were not impacted for any sub-population.

**Fig 7.**
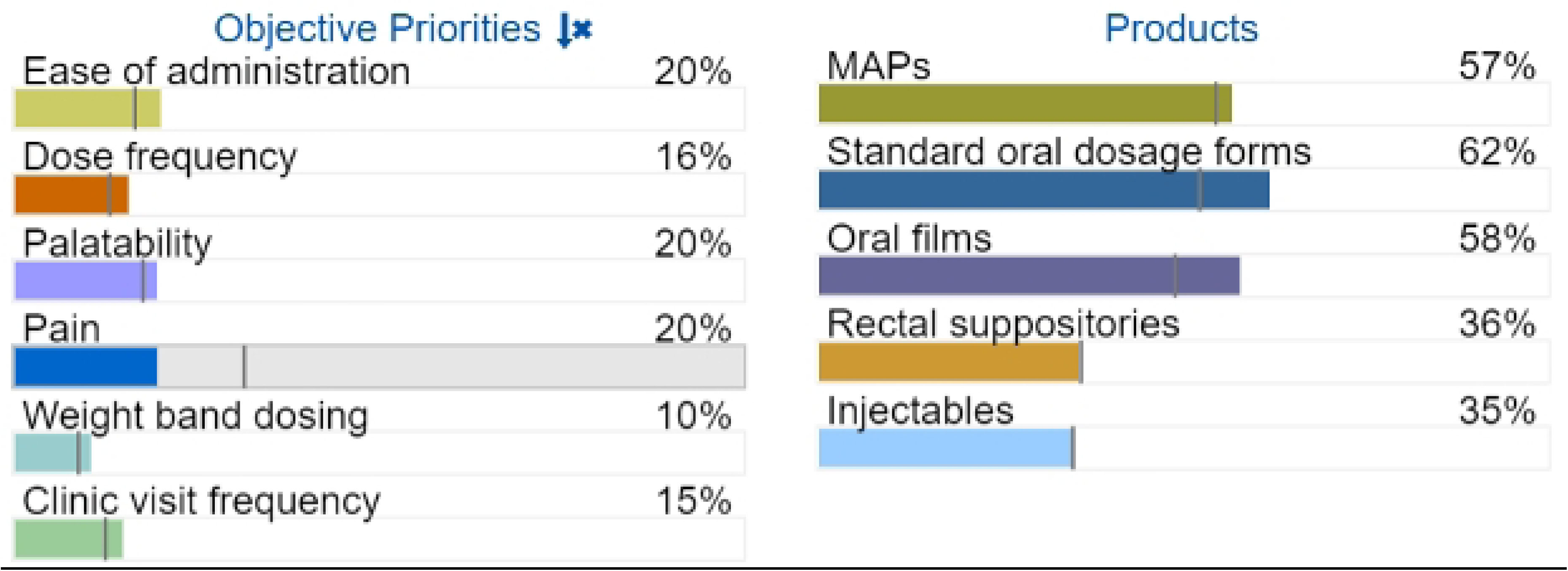
Illustrative example of sensitivity analysis with ease of administration, palatability, and pain being equalized for older children age group.

The aggregation of results allows the highest performing products to emerge across the different age ranges based on their ability to satisfy the product attributes. Table 2 illustrates a summary of the highest ranked options for the pediatric antibiotics case study, where SODFs, MAPS, and oral films emerged as the top three products across the three patient sub-populations. There was significant separation between these top three products and the two lowest scoring products, which were rectal suppositories and injectables. The number next to the product represents the unnormalized relative priority score. SODFs were the first ranked product for one of three pediatric sub-populations, however the separation between the first and third ranked products across all age groups is small (<5.6%).

**Table 2.** Summary of top three products for each pediatric sub-population across all respondents for the AHP pediatric antibiotics exercise.

| Pediatric sub-population | 1<sup>st</sup> ranked product | 2<sup>nd</sup> ranked product | 3<sup>rd</sup> ranked product |
| --- | --- | --- | --- |
| **Neonates and infants (0 up to 2 years)** | SODFs (64.8%) | Oral films (62.2%) | MAPs (61.7%) |
| **Younger children (2 years up to 5 years)** | MAPs (58.1%) | SODFs (53.9%) | Oral films (53.3%) |
| **Older children (5 years up to 12 years)** | MAPs (54.5%) | SODFs (52.3%) | Oral films (48.9%) |

Therefore, it can be concluded that the prioritization of the top three products is equivalent across the age groups.

### Recommendations

The use of AHP in this prioritization exercise highlighted conclusions regarding the product and product attribute priorities that may not have been uncovered without the use of a decision science framework. Based on the AHP survey results, there is a lack of separation between SODFs and the top scoring innovative products, MAPs and oral films, across the three pediatric sub-populations. Although these results suggest that there may be some benefits of MAPs and oral films to meet desired product requirements, current potency limitations for the three APIs remain and warrant pharmacokinetic modeling to establish dosing for pediatric weight bands and determine technical viability. For this reason and considering the small separation of MAPs and oral films from SODFs in the AHP survey, one key finding is that unless there is a strong business case, there is not enough justification to prioritize the large investment required for these innovative technologies over the modest investments that could improve SODFs.

The AHP survey results also highlighted that ease of administration is by far the most important desired product attribute for the youngest pediatric population, and therefore even a minor improvement to administration for this age category known to struggle with oral administration would vastly improve acceptability even though SODFs scored well. Since neonates and infants are not reasonably expected to be able to swallow whole or chewable tablets, future investments should be streamlined towards the development of dispersible tablets in appropriate doses for these three APIs where they do not exist for infants and neonates. While there is are not approved AZM and NFT dispersible tablet formulations for infants, there is a powder for oral suspension available for ACA with dosing for infants (AUGMENTIN trademarked in the US and generic clavulanate potassium equivalents) with United States, United Kingdom and European Union approval [54]. Investments should be made into further assessing existing ACA powder for oral suspension products to determine whether the introduction of a dispersible tablet formulation would increase acceptability among end-users.

Palatability was considered one of the most important desired product attributes, and pooled survey results for each age category indicate that palatability is the biggest shortcoming for SODFs. Although definitive conclusions cannot be made due to the small sample size, the AHP survey results also suggest that improving palatability would have the greatest impact for increased acceptability of SODFs across all pediatric sub-populations. Increased investment by donors in the development of effective taste-masking technologies is strongly recommended to increase acceptability of these medications among pediatric patients.

## Case study 2: Prioritization of pediatric HIV formulations for LMICs

### Objectives and scope

In response to a call to action by WHO, a mapping exercise leveraging AHP methodology was carried out with the goal of assessing compatibility of API-tech pairs and the likelihood of product development success. The APIs, products, and product attributes that were used as the AHP framework inputs for the pediatric HIV treatment and prophylaxis exercise are in Table 3. This scope was defined based on WHO’s PADO5 report outputs [55], technical assessments, and core team engagements prior to initiating the AHP assessment.

**Table 3.** Summary of APIs (top left), products (top right), and product attributes (bottom) in scope for the AHP pediatric HIV exercise.

| APIs | Products |
| --- | --- |
| Cabotegravir (CAB, GSK1265744) | Implants (solid and depot) |
| Dolutegravir (DTG) | Injectables (subcutaneous (SM) and intramuscular (IM)) |
| Islatravir (ISL, MK-8591) | Microarray Patches (MAPs) |
| Lenacapavir (LEN, GS-6207) | Oral Films |
| Tenofovir Alafenamide Fumarate (TAF) |  |
| Product Attributes |  |
| <u>Ease of administration</u>: Platform is easily administered; |  |
| <u>Dose frequency</u>: Platform has an acceptable frequency of dosing (duration of time before the next dose is administered); |  |
| <u>Reversibility</u>: Platform is easily reversible/easily removable; |  |
| <u>Discreetness</u>: Platform is discreet; |  |
| <u>Pain</u>: Platform application is not painful; |  |
| <u>Suitable for pediatric sub-population</u>: I would feel comfortable using the platform for a child in the specified pediatric sub-population/age category listed; |  |
| <u>Dose adjustment</u>: Drug dose adjustments are easily managed as child grows. |  |

### AHP model

The Expert Choice survey was divided into the same two sections as described in the AHP pediatric antibiotics case study section and set up using the same features. The survey was similarly broken into three pediatric sub-populations.

### AHP analysis and results

Of the 133 global stakeholders invited to participate in the pediatric HIV survey, 54 people responded across community member, clinical, and programmatic stakeholder groups. In addition to receiving responses through the online AHP survey, community members and advocates completed a paper-based version of the survey during an in-person Community Advisory Board (CAB) meeting. This ensured participation from a diverse range of community members from over 15 countries in Africa who otherwise may not have access to reliable internet to complete an online survey and allowed for the team to collect responses from non-English speaking CAB members. The CAB members were divided into three sub-groups to align with the three different pediatric sub-populations.

The results were not pooled due to the vastly different responses from the different stakeholder groups. As mentioned in the previous case study, the Expert Choice software has the capability to segment data according to demographic parameters defined by the user. Fig 8 conveys an example of how the Expert Choice software’s visualization of the segmented data.

**Fig 8.**
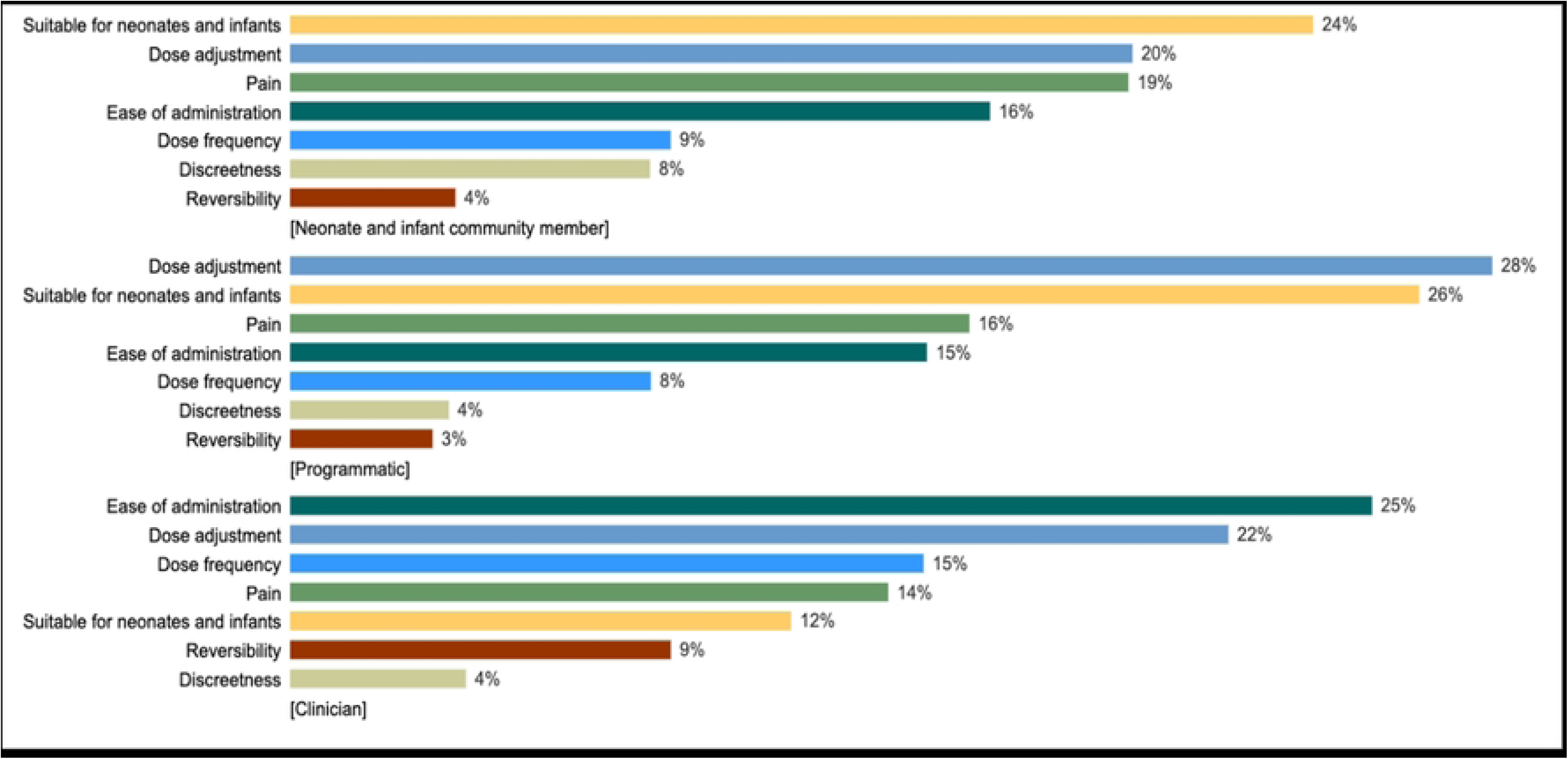
Example of how the Expert Choice results for the relative importance of each desired product attribute can be segmented by stakeholder group within a pediatric sub-population. The top group is neonatal and infant community members, the middle group is programmatic experts, and the bottom group is clinicians.

Aside from the segmentation of the data by stakeholder group, analyses were carried out in a similar manner to the previous case study. Sensitivity analyses were performed and did not yield any significant changes to the outcomes. A summary of the prioritized platforms across the stakeholder groups for each pediatric subpopulation is presented in Table 4. Oral films, injectables and MAPs were repeatedly the top scoring platforms across all pediatric sub-populations and stakeholder groups. There was significant separation between these top scoring platforms and implants.

**Table 4.** Summary of top three platforms for each pediatric sub-population across each stakeholder group for the AHP pediatric HIV exercise.

| Sub-population | Stakeholder group |  |  |
| --- | --- | --- | --- |
|  | Community | Programmatic | Clinical |
| Neonates and infants | Oral film (21%) | Oral film (21%) | Oral film (21%) |
|  | Injection-SC (18%) | MAP-removable (20%) | MAP-removable (18%) |
|  | Injection-IM (16%) | MAP-irremovable (16%) | MAP-irremovable (16%) |
| Younger children | Injection-IM (17%) | Oral film (19%) | Oral film (19%) |
|  | Oral film (16%) | MAP-removable (18%) | MAP-removable (18%) |
|  | Injection-SC (16%) | MAP-irremovable (17%) | MAP-irremovable (17%) |
| Older children | Injection-SC (18%) | Implant-depot (16%) | MAP-removable (16%) |
|  | Injection-IM (17%) | Injection-SC (15%) | Oral film (16%) |
|  | Implant-depot (16%) | Injection-IM (15%) | MAP-irremovable (14%) |

### Recommendations

Like the previous case study, the use of AHP in this prioritization exercise highlighted product preferences across stakeholder groups. Based on the AHP survey results, oral films ranked highly across most stakeholder groups within the three pediatric sub-populations. Despite these stakeholder preferences, API potency limitations remain for oral films. Investment areas for increasing the likelihood of technical success for oral films should focus on optimizing the bioavailability, such as prodrug formulations or increasing the particle surface area by micronization, and effective taste-masking, such as bitter blockers, for the prioritized APIs. Even if bioavailability concerns are addressed, the potential for transformative impact for long-term regimens is limited since oral films cannot be a long-acting product and would instead need be dosed daily.

The AHP framework, specifically the Expert Choice software, allowed the data to be segmented by stakeholder groups. The data segmentation elucidated differences between the preferences of community members and researchers, ultimately highlighting the importance of including community member perspectives in these exercises. For example, there was significant interest in MAPs from the clinical and programmatic experts, however community members did not share the same level of interest in MAP platforms. Despite limited interest by community members, which may be attributed to lack of experience with MAPs, there is current research on the pairing of CAB and TAF with MAP platforms [56–58]. Considering the available scientific evidence on MAP formulations for some of the prioritized APIs, and the known advantages of MAPs such as the removal of cold chain storage, avoidance of gut microbiome changes, and dose flexibility, there is potential for transformative impact if community preferences are met. Initial investments should focus on developing functional MAP prototypes that can be leveraged for end-user studies to evaluate interest and acceptability by users. It is recommended to focus on LEN and ISL MAP prototypes considering the lack of research on these APIs for MAPs, and the potential for these API-tech pairings to succeed according to technical experts consulted during earlier stages of the project.

Both SC and IM injectables were also prioritized across many pediatric sub-populations and stakeholders. There are CAB-LA injectable products approved for use in the United States for HIV prevention and treatment in adults [59, 60], with pediatric dosing under development by the innovator (ViiV Healthcare). There is also a LEN injectable product approved for use in the United States in adult populations [61], however a pediatric formulation has not yet been realized. Near-term investments should focus on formulating a DTG injectable product and modeling to establish dosing for DTG and LEN in pediatric weight bands.

The AHP survey results revealed that implant-depot ranked highly across older pediatric patients, while implant-rod ranked among the lowest prioritized products across all the pediatric sub-populations and stakeholder groups. There is a near-term investment opportunity to further prioritize and then develop implant-depot formulations for these APIs for older children, along with funding the studies required to support these products.

## Discussion

### Lessons learned from the case studies

Multiple lessons can be drawn from the case studies on the power and utility of AHP when applied to complex global health topics. First, the case study findings suggest that AHP methodology is an effective approach for prioritizing products across distinctly different disease areas. Despite differences in the use cases for pediatric antibiotics and HIV products, such as the regimen duration, the findings from both exercises led to the development of product-specific road maps and calls to action. Although a single clear winner did not emerge from the AHP results in either case study, the lack of separation between the top options still added significant value to the recommendations for optimizing investments. Overall, these case studies successfully demonstrate how AHP can be used to prioritize products for research and development investment and provide proof of concept for the exploration of other potential global health applications for AHP.

The case studies also highlight that the Expert Choice software is a powerful tool because it allows a professionally and geographically diverse population to provide inputs in a cost-effective manner. Across both case studies, respondents represented a wide range of technical, clinical, regulatory, and patient perspectives and completed the survey from wherever they were based. In a time where travel funding in global health is under heavy scrutiny due to budget cuts and carbon emission reduction initiatives, it is becoming increasingly more difficult to justify in-person convenings of key stakeholders for strategic discussions. Online AHP tools have the potential to bridge this gap to support systematic, evidence-based decision-making by global health stakeholders across multiple time zones.

WHO has incorporated the outcomes from these case studies into Target Product Profile (TPP) exercises, which reinforces the potential for AHP methods to guide global health strategies, recommendations, and investments. For example, the recommendation to focus on dispersible tablets from the pediatric antibiotics AHP exercise was incorporated into WHO’s final TPP for pediatric formulations of azithromycin and nitrofurantoin [62]. The adoption of these recommendations into WHO TPPs emphasizes the ability for AHP exercise outputs to be incorporated into normative documents that will be used by product developers and funders to inform research and development agendas.

### Limitations

There are limitations to the outcomes of the case studies, which reiterate the broader limitations of AHP. For instance, the sample size of both case studies was small. The aggregated response rate across both case studies was 38% despite multiple attempts to get diverse and individual responses. Although the team was able to generate action-based roadmaps from the results, statistically significant conclusions could not be made. The low response rates may be attributed to multiple factors such as the survey length and complexity, which is inherent to a survey involving AHP pairwise comparisons. Additionally, global health professionals are oftentimes juggling multiple paid and unpaid roles, and as a result experts may have been unable to respond due to busy schedules. Creative engagement and follow-up strategies should be explored in future global health related AHP exercises to encourage a larger number of responses. For example, global advocacy groups should be leveraged where appropriate to gather responses from patients and caregivers that take or administer the medications being considered. Additional approaches should also be explored to reduce time requirements, such as deploying a multi-phased survey approach or further modifying the Expert Choice software settings to reduce the number of judgements options.

AHP models rely on critical thought and focused effort to define the inputs and as a result there is always a risk of important criteria or alternatives being overlooked. Even if all criteria and alternatives were identified it is not realistic to capture all potential inputs, especially if the survey is maintained at a manageable length. During the case studies, a small team of subject matter experts was assembled with technical, clinical, regulatory, and programmatic backgrounds to define and prioritize survey inputs.

Although there will always be some level of unknowns or external factors that are not accounted for in an AHP exercise, the assembly of a multidisciplinary team to determine the inputs can ensure that the AHP survey outcomes address the highest priorities.

Another limitation of AHP, which is also a learning, is that the tool itself is not designed to replace the decision maker’s thinking. The primary goal of all decision science tools and frameworks is to facilitate discussions and better organize the thoughts of decision makers while making their rationale more presentable to others. Thus, successful utilization of AHP still depends on adequate time and resources to carry out the process and interpret the findings to support decision making. Even though an upfront investment is necessary for AHP in the short-term, it can yield cost and time savings in the long term if well-executed.

### Additional opportunities to apply AHP to global health

Although the case studies were focused on applying AHP to prioritize investments for life saving medicines, the successful utilization of AHP methodology in this context suggests that there is a broader opportunity to leverage AHP to prioritize global health investments. For instance, AHP could be used by donors to prioritize the scope of their portfolios and support the awardee selection process. The framework could also be used by organizations to evaluate strategic partnership options and inform partner selection according to the objectives of the program. There is also an opportunity to embed AHP methods into operational research activities. Although there is a wide range of literature on how operational research techniques can be applied to global health topics, AHP specific examples are limited [63]. Integrating AHP methods into operational research studies could uncover additional priorities or considerations that may not have otherwise been identified.

The current emphasis on local and regional manufacturing in LMICs is a compelling example of where there could be a significant benefit to breaking down a large, multi-faceted goal into smaller decisions using AHP. The COVID-19 pandemic put a spotlight on the dramatic inequities in accessing health commodities between high- and low-income settings and as a result there has been increased interest in regional manufacturing among influential global health donors such as Unitaid, PEPFAR, and The Global Fund. For example, Unitaid published a position paper on local manufacturing [64] and also recently posted an Expression of Interest [65] for strengthening sustainable manufacturing of therapeutics for maternal health, malaria, and HIV. WHO has also been actively supporting regional manufacturing initiatives through their Local Production and Assistance Unit [66], as well as through hosting three World Local Production Forums in 2021, 2023, and 2025 [67]. The uptick of local and regional manufacturing initiatives amidst an uncertain and constrained funding landscape presents a strong opportunity to utilize AHP to systematically prioritize potential areas for investment according to impact-driven requirements. AHP would serve as a powerful tool for enabling a wide range of stakeholders to define criteria for impact of a regional manufacturing initiative and evaluate the performance of potential interventions against those criteria based on their level of importance. AHP could also be used to prioritize products or product categories to pursue under local or regional manufacturing initiatives, allowing for a quantitative and qualitative assessment according to factors such as disease burden, complexity of the manufacturing process, product risk classification, existing capacity, and product availability, among others. Other applications of AHP to local and regional manufacturing could include prioritization of countries or industries. In a time where the next global public health emergency could be around the corner and public healthcare budgets are being strained more than ever, AHP could help catalyze local and regional manufacturing investments with the greatest chance of far-reaching impact.

### Conclusion

Although AHP is widely used across many industries, it has been underutilized in LMIC based health applications. Two AHP surveys were built, disseminated, and analyzed using Expert Choice software with the goal of prioritizing pediatric antibiotics and HIV formulations for LMICs. Both case studies resulted in recommendations for how to prioritize investments and ultimately advance development of the most promising API-tech pairs based on their ability to meet the highest priority user requirements, which were positively received by the WHO and other key stakeholders. The encouraging results from these case studies suggest that there is an opportunity for the global health community to embrace AHP methods across a variety of use cases. Considering the turbulent global health funding landscape, the broader uptake of AHP by global health partners could improve efficiencies in the allocation of resources, prioritization of investments, and implementation of programs.

## Data Availability

The data for this manuscript can be accessed in the attached Supporting Information file. See S1 Dataset.

## Acknowledgments

We thank all the individuals that participated in the pediatric antibiotic and HIV case studies, including our steering group members in the pediatric antibiotic and HIV case studies, Jason Brophy, Andy Carmone, Chipo Chimhundu, Sheetal Ghelani, Herb Harwell, Nagalingeswaran Kumarasamy, Linda Lewis, John Miller, Brian Nzano, Dani Resar, Pablo Rojo and George Siberry We also thank our WHO and GAP-f collaborators, Martina Penazzato, Asma Hafiz, and Bernadette Cappello, for their support.

